# Differential Methylation at Novel Putative Imprinting Control Regions on Chromosome 20 Associated with Childhood Obesity

**DOI:** 10.64898/2026.09.02.26362065

**Authors:** Catherine F. Everly, Terrence K. Allen, David A. Skaar, Bruce A. Corliss, Dereje D. Jima, Rachel L. Maguire, Halah Jadallah, Randy L. Jirtle, Susan K. Murphy, Jung-Ying Tzeng, Cathrine Hoyo

## Abstract

DNA methylation may link early-life exposures to later obesity risk; however, most studies examine individual CpG sites using commercial arrays that cover <5% of the genome and often overlook biologically-relevant regulatory regions. Imprinting control regions (ICRs) regulate parent-of-origin gene expression and are enriched for growth and metabolic genes, representing strong candidates for developmental programming of obesity. In 586 participants of the Newborn Epigenetics STudy (NEST), we leveraged the novel Human Imprintome Array to measure methylation at >1,000 characterized and putative ICRs genome-wide in umbilical cord blood. We applied principal component and kernel machine regression to identify ICR methylation at birth associated with sustained childhood obesity, defined as BMI ≥95^th^ percentile during most of childhood. Sustained childhood obesity was associated with differential methylation at seven ICRs in cord blood, with 2-10% mean differences between obesity and normal weight groups, including novel regions localized to chromosome 20q11.1-20q11.21 (ICR_1182, ICR_1179, ICR_1181, ICR_1177, ICR_1180, and ICR_1165), mapping to or near *CDC27P4, LINC01597, DUX4L37,* and *DUX4L34,* and ICR_927 mapping to previously characterized *ZNF597/NAA60.* Most ICRs overlapped multiple transcriptional regulator binding sites. Associations persisted in peripheral blood collected in later childhood at ages 8-16 years. Among offspring of mothers with pre-pregnancy obesity, an additional 50 associations were identified. Sex-stratified analyses revealed no further insights. These findings identify novel ICRs—particularly on chromosome 20q—as potential early-life epigenetic biomarkers of childhood obesity. If replicated, these stable ICR methylation patterns may identify children at risk for obesity at birth, informing targeted prevention before obesity develops and guiding novel therapeutic strategies.

## Introduction

Childhood obesity is a growing health crisis worldwide. In the United States, obesity prevalence among youth aged 2-19 years old was 19.8% in 2024, but this prevalence varies substantially by socio-economic status and ethnicity (1–3). Once considered a health crisis only in high-income countries, childhood obesity rates have also surged in low- and middle-income countries during the late 20^th^ to early 21^st^ century (4). Childhood obesity is a strong predictor of adult obesity, with obese youth around 5 times more likely to maintain that status into adulthood compared to non-obese peers (5–7). Obesity is a major risk factor for cardiovascular disease, certain cancers, type 2 diabetes, and some neurological disorders (8–12), with all-cause mortality rates from these diseases also associated with childhood obesity (10,13–15). In addition, children with obesity face psychological harm from low self-esteem, depression, and discrimination by their peers (16). Behavioral interventions focused on caloric intake and exercise are difficult to maintain long-term (17,18), and glucagon-like peptide-1 receptor agonists (GLP-1RAs), while promising and approved to treat obesity in children, are currently limited by age restrictions, cost, and uncertainty of the unintended and long-term health consequences in pediatric populations (19–22). Because childhood growth occurs at varying rates of height gain and adiposity accumulation (23,24), there is an urgent need for early-life biomarkers that can identify children on trajectories toward sustained obesity before these patterns become clinically evident.

Mounting evidence supports that environmental exposures during early development can alter epigenetic regulation and contribute to obesity risk in children. Consistent with the Developmental Origins of Health and Disease (DOHaD) hypothesis (25), periconceptional and prenatal exposures—including maternal stress, maternal smoking, maternal obesity, paternal obesity, and environmental pollutants—can induce epigenetic changes during prenatal development (26–31), with CpG site methylation being the most extensively studied. Differential methylation at multiple CpG sites has been associated with childhood obesity (32,33), including at regulatory regions of known obesity genes, such as the promoter for leptin (*LEP*), which regulates energy balance, appetite, and metabolism (34–36). Genome-scale interrogation of differential methylation using the Illumina Infinium HumanMethylation450 (450K) or MethylationEPIC (EPIC) BeadChip arrays has enabled the discovery of novel loci and pathways; however, these findings are not always reproducible. One major challenge to reproducibility is that genome-scale methylation studies often interrogate CpG sites that remain environmentally responsive and are tissue-specific throughout the life course (37). Consequently, results may differ across studies depending on tissue type and timing of sample collection (38). Furthermore, obesity is often measured at a single time point, despite the nonlinear nature of childhood growth, such that associations may vary depending on when adiposity is assessed. Evidence suggests that the association between childhood obesity and poorer cardiometabolic health is stronger and more consistent when obesity is sustained over time (39,40). Finally, the genome coverage of these arrays is low (< 5% of CpG sites in the human genome), leaving large regions of the genome unexplored. Together, these limitations highlight the need to identify epigenetic marks with known developmental stability and to study obesity as a sustained longitudinal phenotype, given the dynamic nature of the epigenome and the nonlinear growth patterns characteristic of childhood.

One approach to address these challenges is to focus on DNA methylation at genomic regions whose stability across development is established and whose regulatory roles suggest they precede disease onset. Imprinting control regions (ICRs) are CpG-dense regulatory elements that establish and maintain parent-of-origin–specific monoallelic gene expression of imprinted genes through differential allelic DNA methylation that silences one parental allele (41–43). Cytosine methylation at ICRs is established in the germline and normally resists the global epigenetic postfertilization methylation reprogramming that takes place in the early embryo (44–46). Thus, ICR methylation patterns are already set prior to germ layer specification and are maintained with high fidelity during tissue differentiation in the developing embryo. This early establishment, combined with the mitotic heritability of DNA methylation, results in ICR methylation being highly similar across tissues, including in DNA obtained from accessible tissues such as blood (47,48). These methylation marks are critical for regulating dosage of essential developmental imprinted genes, and are maintained throughout an individual’s life (47–49), and aberrant ICR methylation is associated with metabolic, growth, and neurological disorders, including severely disabling conditions (50–52). This spatial and temporal methylation stability of ICRs makes them attractive targets for biomarker discovery for the early identification of obesity susceptibility.

Despite their importance, only 24 ICRs regulating ∼200 genes had been characterized until recently (53). With recent advances in whole genome sequencing and computational capacity, over 1,000 novel putative ICRs have been identified (54). These putative ICRs were predicted using stringent bioinformatic analyses of whole-genome bisulfate sequencing data. However, unlike experimentally validated ICRs, their role in regulating genomic imprinting and the establishment of allele-specific methylation patterns in somatic tissues have not yet been experimentally confirmed. Cytosine methylation at these regions can be reproducibly quantified using the newly developed Human Imprintome Array, which targets > 70% of characterized and newly identified putative ICRs, compared with only 10.5% coverage by the EPICv1 array (55). Herein we report the identification of ICRs in cord blood obtained at birth that are associated with sustained childhood obesity, leveraging longitudinal BMI anthropomorphic measures and imprintome-wide profiling to evaluate persistence of cord blood DNA methylation patterns with those obtained in peripheral blood during childhood and adolescence.

## Materials and Methods

### Participant Information

Participants in this study were mother-child pairs enrolled in the Newborn Epigenetic STudy (NEST) between 2005 and 2011 in Durham, North Carolina, USA. Enrollment protocols for the NEST cohort are detailed elsewhere (56,57). Briefly, the study inclusion criteria were pregnant women who were at least 18 years of age, English- or Spanish-speaking, and intending to deliver at Duke University Hospital or Duke Regional Hospital to enable collection of umbilical cord blood specimens. At enrollment, mothers had a median gestational age of 11-12 weeks (range 6-27 weeks). A total of 2,681 mother-child pairs were enrolled in NEST, and umbilical cord blood for DNA methylation analysis was successfully collected from over two-thirds of the children at delivery. Women were further excluded after enrollment if they experienced fetal death, withdrew from the study, or did not respond to further communication. Information on maternal health, demographics, and other pre-pregnancy characteristics was collected at enrollment and at birth.

Of the 2,404 live births, 1,454 children after age 2 years were followed at least three times for height and weight measurements to assess consistency in their body mass index (BMI) measurements. The median interval between measurements was 63 days (interquartile range = 20-196). After excluding those with biologically implausible BMI values and children born extremely preterm, 1,442 children remained. Among these children, 691 also had available umbilical cord blood DNA methylation data. After further excluding those with incomplete covariate data or DNA methylation data that failed QC, the final analytical sample consisted of 586 mother-child pairs (Supplementary Figure S1). Across all study visits, BMI measurements were collected when children were between 2 and 16 years of age.

Those excluded were compared to the 586 included participants with respect to maternal and infant characteristics using Chi-Square tests (for categorical variables) and t-tests (for continuous variables). Most characteristics were not statistically different (p > 0.05). Significant differences were observed for maternal race/ethnicity, maternal age at delivery, gestational age at delivery, and infant birthweight (p < 0.05); however, the differences corresponded to small standardized mean differences of | < 0.35| (Supplementary Table S1).

### Body composition categorization of participating children

After birth, the children were followed longitudinally throughout childhood and adolescence to obtain anthropometric measurements, including height and weight. These measurements were obtained during scheduled NEST study visits or routine doctor’s visits using a Tanita stadiometer (Tanita Corporation, Tokyo, Japan). BMI was calculated using the SAS Program for CDC Growth Charts, which accounts for the child’s height, weight, age, and sex (58). Obesity was defined according to CDC guidelines as BMI at or above the 95^th^ percentile for a child’s age and sex. To expand on prior studies that largely estimated childhood obesity based on a single BMI measurement, we focused on sustained longitudinal obesity, defined as having a BMI > 95^th^ percentile for at least 80% of observations throughout childhood. This approach is consistent with a recent publication that assigned children to the BMI category present in at least 50% of measurements across three visits (59). Although trajectory modeling approaches have been widely used to characterize longitudinal patterns of childhood BMI (60,61), we selected a classification approach to maximize clinical interpretability and reproducibility, as these categories may be readily applied to any cohort with longitudinal BMI data without requiring cohort-specific statistical modeling.

Children were categorized into three body composition groups: normal weight, sustained obesity, and intermittent obesity (those obese < 80% of the time). The 80% cutoff is intended to account for children whose BMI oscillated over time and to ensure they are categorized based on where the *majority* of their measurements fall. Children categorized as normal weight (n = 417) were those with at least 80% of their BMI measurements below the 95^th^ percentile. Although referred to as the normal weight group, some children may have had individual BMI measurements in the CDC-defined overweight range because classification was based on longitudinal BMI patterns rather than on each cross-sectional BMI measurement. Children with sustained obesity (n = 61) were those with at least 80% of their BMI measurements at or above the 95^th^ percentile. Among children with at least 80% of their BMI measurements in a single category, BMI measurement counts ranged from 5 to 118 (mean = 21.2, standard deviation = 14.5). The rest of the children were categorized as “intermittent obesity,” i.e. obese < 80% of the time (n = 108). This category included children who measured a mixture of obese and normal weight measurements and are therefore at an elevated risk of developing obesity than children consistently normal weight (7). Approximately half of the intermittently obese children demonstrated increasing BMI over time and transitioned from normal weight to obese measurements, indicating disproportionate weight gain relative to height over time.

### Covariate data

Covariate selection was guided by 1) prior literature on maternal and early-life factors associated with childhood obesity, and 2) known relationships between those factors and differential DNA methylation. Variables considered were maternal smoking, maternal race/ethnicity, maternal education, maternal obesity, parity, maternal age at delivery, gestational age at delivery, infant birthweight, infant sex, breastfeeding status, and delivery route. Maternal smoking, maternal race/ethnicity, and maternal obesity alter methylation of known ICRs and other CpG sites (31,50,62–64). Recent meta-analyses of 450K/EPIC data also suggest maternal education—likely a proxy for socio-economic status—is associated with differential CpG methylation, as is offspring sex (65,66). Breastfeeding status has been linked to childhood obesity risk, and serves as an approximation of early childhood nutritional status and eating behaviors (67–69). Child birthweight was not included as a covariate in downstream analyses because it lies on the causal pathway between prenatal exposure (e.g. maternal obesity) and childhood adiposity. Prenatal risk factor data were obtained at enrollment using a standardized questionnaire, parturition data were abstracted from medical records, and postnatal data were collected in the clinic or during home visits.

### DNA methylation data collection and processing

DNA was extracted from umbilical cord blood collected at birth and from peripheral blood collected during one follow-up visit using PURGENE reagents (QIAGEN N.V.). DNA methylation analysis was done at TruDiagnostic, Inc., using the Human Imprintome Array Bead Chip. Specifically, DNA (500 ng) was bisulfite-converted, assigned to chip wells on the Illumina Human Imprintome Array Bead Chip, amplified, hybridized to the array, stained, washed, and imaged with an Illumina iScan SQ instrument to obtain raw image intensities. Sample processing followed the standard Illumina Infinium protocols and instrumentation used for the HumanMethylation450K and EPIC BeadChip platforms; however, all study samples were hybridized to the Human Imprintome Array BeadChip rather than the HumanMethylation450K or EPIC arrays. To preprocess DNA methylation values, the *SeSAMe* package was used due to its compatibility with Illumina custom arrays (70). The TruDiagnostic Human Imprintome Array (*tdhia*) R package was used to implement the *SeSAMe* preprocessing workflow for Human Imprintome Array data (71). We used the *openSesame* and *getBetas* functions to generate a beta-value matrix for the probes and a corresponding SeSAMe signal detection p-value matrix, which is used for quality control. Raw channel signal intensities were normalized using non-linear dye bias correction, and background subtraction was applied using the NOOB method (normal-exponential out-of-band signal), which deconvolutes out-of-band fluorescent signal from true signal (72). Quality control filtering was implemented at the probe and patient level. For umbilical cord blood data, probes were removed from analysis if more than 10% of their measurements had signal detection p > 0.20. For the data from peripheral blood collected later in life, the maximum probe fail rate was increased from 10% to 15% to ensure that the majority of probes in the original analysis were present for analysis at the later timepoint. For both umbilical cord and peripheral blood, children were removed from analysis if greater than 5% of their measurements had p > 0.20. DNA methylation was calculated as beta-values for each CpG site by taking the ratio of the intensity of methylated bead types to the total intensity of methylated and unmethylated bead types. As a result, beta-values are continuous values of 0–1, where 0 reflects 0% methylated or fully unmethylated, and 1 reflects 100% methylation or fully methylated at the CpG site. Any CpGs that did not map to a unique genomic position or an ICR were discarded. The final CpG beta matrix used in the analysis contains 6,711 CpG sites, corresponding to 1,022 ICRs.

### Statistical analysis

All statistical analyses were conducted in R version 4.5.1. (73), using the tdhia R package (71) and custom analysis scripts available in the publicly accessible hoyo_published_analyses GitHub repository.

### Association of maternal and child characteristics with childhood obesity

To determine if covariates were associated with sustained or intermittent obesity status in children and thus warrant inclusion in our multivariate models, maternal and child characteristics were tested individually for their marginal association with the obesity body composition categories using logistic regression.

### Association of ICR methylation with childhood obesity

To identify ICRs associated with childhood obesity, we conducted CpG- and ICR-level association analyses comparing normal weight children (i.e. the reference group) to children with sustained or intermittent obesity. Firstly, despite the dependence among CpG sites within an ICR, we built logistic regression models to individually test the association of each CpG and covariates to each obesity outcome. Secondly, for ICR-level analyses, we used principal component (PC) regression and kernel machine (KM) regression, which offer complementary perspectives to assess the joint effects of potentially correlated CpG sites within an ICR. PC regression summarizes methylation across an ICR using the top PCs that explain the majority of the total CpG variance and evaluates their joint effect with child obesity in logistic regression models. PCs are constructed unsupervised, based solely on the predictor data. KM regression assesses the aggregate effects of all CpGs within an ICR by modeling their combined contribution to obesity risk through a linear kernel function.

### CpG-level Analysis

Similar to 450K/EPIC array analysis, we assessed the association between CpG methylation and sustained or intermittent obesity using logistic regression, adjusting for maternal smoking, maternal education, maternal race/ethnicity, maternal obesity, breastfeeding status, and sex of the child. We also conducted stratified analyses by child sex and maternal pre-pregnancy obesity, since these factors are likely to act as effect modifiers. To account for multiple testing across 6,711 CpG sites, we controlled the false discovery rate (FDR) using the q-value method implemented in the qvalue R package, with a q-value < 0.05 as the significant threshold and a q-value < 0.20 as a less conservative, exploratory cutoff.

### ICR-level Analysis

In PC regression, first we standardized the methylation values at each CpG site to ensure comparability across CpGs. Next, we obtained the top PCs of standardized CpG methylation data in an ICR accounting for at least 80% of the total methylation variance. For ICRs containing only a single CpG site that passed quality control (177 ICRs in total), the original methylation value was used instead of PCs. Next, we performed logistic regression with childhood obesity status as the outcome. For each ICR, we compared a full model including PCs and covariates to a reduced model including covariates only, and used the likelihood ratio test (LRT) to assess the significance of the inclusion of PC terms.

In KM regression, we used the sequencing kernel association test (SKAT) as implemented in the SKAT R package, using a linear kernel and equal weights (74). Similar to the CpG-level analysis, PC and KM regression were implemented for analyses stratified by child sex and maternal pre-pregnancy obesity. The same covariates were adjusted for as in the CpG-level analysis. FDR correction was applied across the 1,022 ICRs using the same q-value-based thresholds as in the CpG-level analyses.

### ICR-level Sensitivity Analysis

The majority of the ICRs are recently predicted putative regions whose boundaries have not yet been experimentally validated. Consequently, some CpGs assigned to an ICR may not belong to the functional regulatory region, while neighboring putative ICRs may ultimately represent a single regulatory domain. PC and KM regression are better at handling possible heterogeneous methylation patterns among CpGs within ICRs than a simple arithmetic mean since they do not assume each CpG contributes equally to the signal. However, in sensitivity analysis, for simplicity, we also determined if the arithmetic mean across CpGs within each putative ICR would perform at identifying associations with childhood obesity. To test this, the CpG beta values were averaged across each ICR for each individual. Next, we assessed the association between average ICR methylation and sustained or intermittent obesity using logistic regression, adjusting for maternal smoking, maternal education, maternal race/ethnicity, maternal obesity, breastfeeding status, and sex of the child. This analysis was run for each stratified and unstratified group. FDR correction was applied across the 1,022 ICRs.

### Stability of ICRs over time

The stability of methylation marks at ICRs is a hallmark feature that distinguishes them from the rest of the genome, as they must be maintained throughout an individual’s life and across individuals to control gene dosage. To evaluate stability, a subset of 100 children had methylation measured on the Human Imprintome Array in both umbilical cord blood collected at birth and again in peripheral blood collected at age 8-16 years. Of these children, 86 were classified as normal weight, and 14 as sustained obesity. Stability was analyzed for (1) within-person longitudinal stability and (2) stability of obesity-associated differences. For both methods, the mean CpG methylation was calculated across each ICR. Within-person longitudinal stability was assessed by calculating the Pearson’s correlation coefficient (r) between birth and late childhood/adolescence. Correlations were calculated separately for the normal weight and sustained obesity groups. To analyze the stability of obesity-associated differences, the mean difference in methylation between normal weight and sustained obesity children was calculated for each ICR at each timepoint. These ICR-level differences were then compared between birth and late childhood/adolescence to determine whether their magnitude and direction were stable over time. In addition, we conducted PC regression on the subset of ICRs found to be significant at birth in the unstratified normal weight versus sustained obesity analysis to evaluate whether ICR-level associations persisted at later childhood/adolescence timepoints. Multiple testing was controlled using the Benjamini-Hochberg (BH) FDR method, as implemented in the p. adjust function in R.

### Functional Analysis

To investigate any possible functional relevance of genes identified as significant across the analyses, we used QIAGEN Ingenuity Pathway Analysis (IPA) (75). IPA provides context for the biological pathways that may mediate the obesity phenotype through methylation at ICRs. All genes mapping to or near the significant CpGs or ICRs with q < 0.05 across all the stratified and unstratified analyses were extracted and input to IPA. The reference gene set consists of all genes mapping to or near CpGs or ICRs represented on the array. IPA Core Analysis was conducted.

## Results

### Characteristics of study participants

Maternal and child characteristics of the 586 mothers and children included in these analyses are summarized in Supplementary Table S2. The prevalence of sustained obesity was 10.4% (n = 61) and that of intermittent obesity was 18.4% (n = 108). Childhood obesity did not significantly vary with parity, maternal age at delivery, gestational age at delivery, breastfeeding status, delivery route, or sex of the child. However, as expected, children of women with pre-pregnancy obesity were significantly more likely to exhibit sustained obesity (β = 1.18, 95% CI: 0.63–1.73; SE = 0.28, p = 2.54 × 10^−5^) or intermittent obesity (β = 1.09, 95% CI: 0.65–1.52; SE = 0.22, p = 9.70 × 10^−7^). Children born to Hispanic mothers were also significantly more likely to exhibit sustained obesity (β = 1.42, 95% CI: 0.67–2.17; SE = 0.38, p = 2.05 × 10^−4^) or intermittent obesity (β = 0.85, 95% CI: 0.28–1.42; SE = 0.29, p = 3.57 × 10^−3^) than those born to Non-Hispanic White mothers. Maternal education, smoking, and birthweight associations were only significantly associated with either sustained obesity or intermittent obesity. Significantly associated covariates, in addition to sex of the children and breastfeeding status, were adjusted for in our CpG- and ICR-level analyses.

### Putative imprinting control regions associated with sustained childhood obesity

The normal weight (n = 417) and sustained obesity (n = 61) groups were compared at the CpG-level using logistic regression. After adjusting for covariates and multiple comparisons, sustained obesity was associated with 11 significant CpGs (q < 0.05), corresponding to five ICRs–one mapping to a known imprinted gene and four novel (Figure 1, Table 1). These ICRs (ICR_1182, ICR_1179, ICR_1181, ICR_1165, and ICR_927) range in size from 212 to 1,039 base pairs and contain 4 to 21 CpG sites. These ICRs map within or near genes *CDC27P4, LINC01597, DUX4L34,* and *ZNF597/NAA60*. ICR_927 overlaps with *ZNF597* and *NAA60,* which are experimentally characterized imprinted genes with maternal expression (76,77). The Manhattan plot in Figure 1a highlights CpGs with q < 0.05.

**Figure 1.**
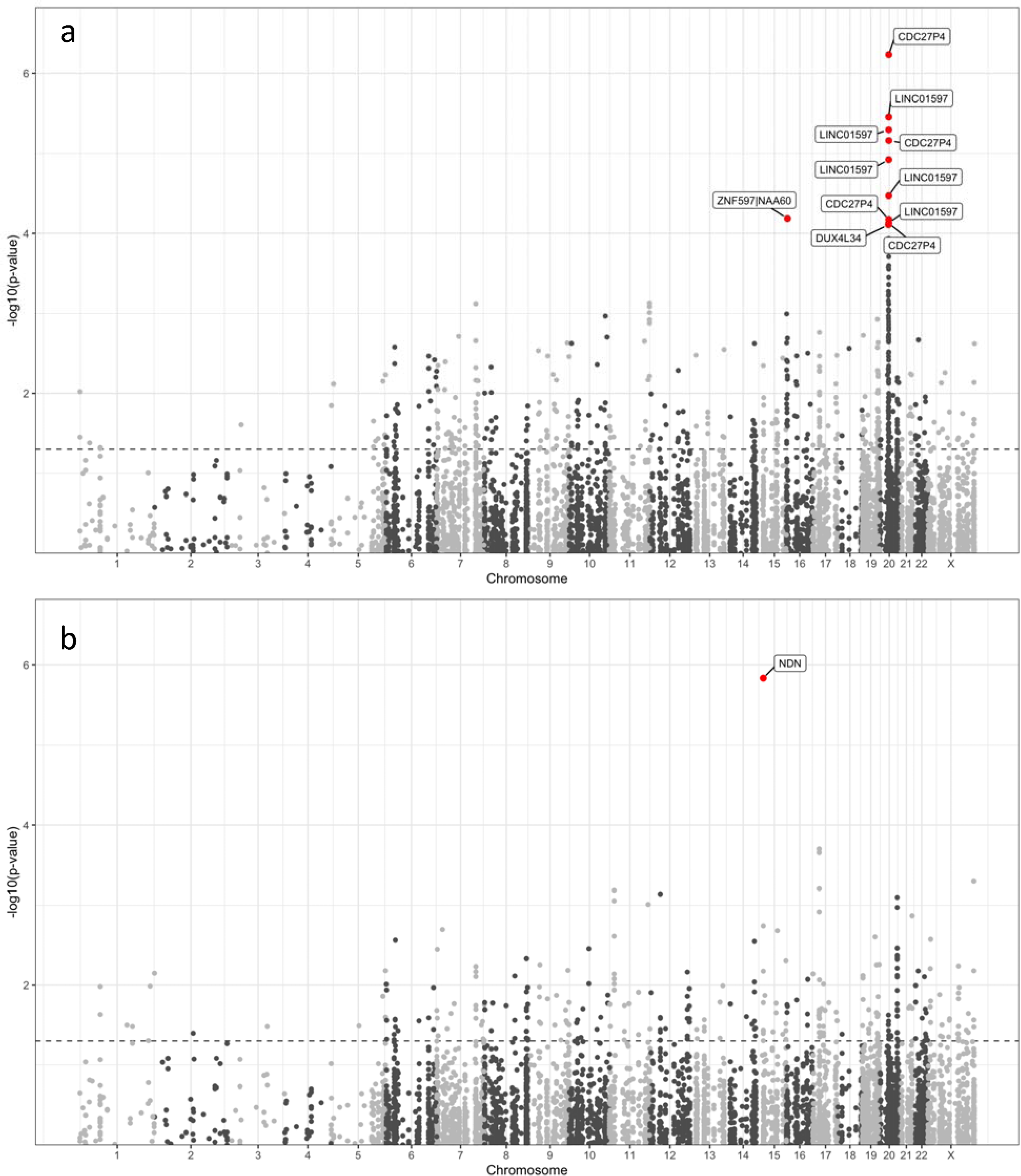
Manhattan plots showing results from CpG-level logistic regression models. The y-axis represents -log10(p-values). Significant CpG sites with q < 0.05 are highlighted in red and labeled with the nearest gene. The horizontal gray dotted line corresponds to unadjusted p = 0.05. (a) Significant CpGs associated with sustained obesity. Four CpGs mapped nearest to *CDC27P4*, five CpGs mapped to or near *LINC01597*, one CpG mapped to *DUX4L34*, and one CpG mapped to *ZNF597/NAA60*. (b) Significant CpGs associated with intermittent obesity. One CpG mapped to *NDN*.

**Table 1.** Significant CpGs and ICRs associated with sustained childhood obesity. All significant CpGs (by logistic regression q < 0.05) or ICRs (by PC regression or KM regression q < 0.05) for the sustained obesity group. The CpG logistic regression estimates and odds ratios are per 1% increase in methylation. Known imprinted genes are denoted with a *. CpG- or ICR- level q < 0.05 are bolded.

| ICR | Nearest Gene | ICR Genomic Coordinates (GRCh38) | Number of CpG sites measured within ICR | CpG | CpG Logistic Regression raw p (q) | CpG Logistic Regression Estimate | CpG Logistic Regression Odds Ratio (95% CI) | Mean methylation difference (percentage points; sustained obesity – normal weight) | PC Regression raw p (q) | KM Regression raw p (q) |
| --- | --- | --- | --- | --- | --- | --- | --- | --- | --- | --- |
| ICR_1182 | CDC27P4 | chr20:30488015-30488659 | 15 | cg25130503 | $5.86 \times 10^{-7}$<br>**(3.65 × 10<sup>-3</sup>)** | 0.06<br>(0.04, 0.09) | 1.06<br>(1.04, 1.09) | 9.71% | $3.96 \times 10^{-6}$<br>**(2.78 × 10<sup>-3</sup>)** | $7.79 \times 10$<br>**(7.97 × 10)** |
| | | | | cg25130505 | $6.88 \times 10^{-6}$<br>**(1.07 × 10<sup>-2</sup>)** | 0.16<br>(0.09, 0.23) | 1.17<br>(1.09, 1.26) | 2.93% | | |
| | | | | cg25130559 | $6.71 \times 10^{-5}$<br>**(4.41 × 10<sup>-2</sup>)** | 0.04<br>(0.02, 0.06) | 1.04<br>(1.02, 1.06) | 9.52% | | |
| | | | | cg25130500 | $7.47 \times 10^{-5}$<br>**(4.41 × 10<sup>-2</sup>)** | 0.04<br>(0.02, 0.05) | 1.04<br>(1.02, 1.06) | 9.52% | | |
| ICR_1179 | LINC01597 | chr20:30283622-30283834 | 7 | cg25128653 | $3.48 \times 10^{-6}$<br>**(1.05 × 10<sup>-2</sup>)** | 0.11<br>(0.06, 0.16) | 1.12<br>(1.07, 1.17) | 4.47% | $1.10 \times 10^{-5}$<br>**(3.87 × 10<sup>-3</sup>)** | $1.73 \times 10$<br>**(8.81 × 10)** |
| | | | | cg25128649 | $5.05 \times 10^{-6}$<br>**(1.05 × 10<sup>-2</sup>)** | 0.25<br>(0.14, 0.35) | 1.28<br>(1.15, 1.42) | 1.86% | | |
| | | | | cg25128648 | $1.20 \times 10^{-5}$<br>**(1.49 × 10<sup>-2</sup>)** | 0.09<br>(0.05, 0.12) | 1.09<br>(1.05, 1.13) | 5.34% | | |
| | | | | cg25128647 | $3.38 \times 10^{-5}$<br>**(3.50 × 10<sup>-2</sup>)** | 0.06<br>(0.03, 0.10) | 1.07<br>(1.03, 1.10) | 6.36% | | |
| ICR_1181 | LINC01597 | chr20:30288471-30289510 | 4 | cg13023119 | $7.40 \times 10^{-5}$<br>**(4.41 × 10<sup>-2</sup>)** | 0.11<br>(0.05, 0.16) | 1.11<br>(1.05, 1.17) | 3.19% | $8.48 \times 10^{-4}$<br>(1.04 × 10<sup>-1</sup>) | $1.45 \times 10$<br>**(3.61 × 10)** |
| ICR_1177 | DUX4L37 | chr20:29878870-29878965 | 3 | NA | NA | NA | NA | NA | $8.87 \times 10^{-4}$<br>(1.04 × 10<sup>-1</sup>) | $1.77 \times 10$<br>**(3.61 × 10)** |
| ICR_1180 | LINC01597 | chr20:30284340-30284720 | 16 | NA | NA | NA | NA | NA | $1.15 \times 10^{-3}$<br>(1.15 × 10<sup>-1</sup>) | $1.60 \times 10$<br>**(3.61 × 10)** |
| ICR_1165 | DUX4L34 | chr20:29411151-29411665 | 21 | cg25121519 | $7.79 \times 10^{-5}$<br>**(4.41 × 10<sup>-2</sup>)** | 0.09<br>(0.05, 0.14) | 1.09<br>(1.05, 1.15) | 3.09% | $8.65 \times 10^{-3}$<br>(2.89 × 10<sup>-1</sup>) | $4.09 \times 10$<br>(6.97 × 10) |
| ICR_927 | ZNF597*\|NAA60* | chr16:3443280-3444094 | 20 | cg21026074 | $6.52 \times 10^{-5}$<br>**(4.41 × 10<sup>-2</sup>)** | 0.10<br>(0.05, 0.14) | 1.10<br>(1.05, 1.15) | 2.94% | $5.48 \times 10^{-4}$<br>(9.60 × 10<sup>-2</sup>) | $1.46 \times 10$<br>(4.65 × 10) |

Accounting for the correlation structure among CpG sites within each ICR using principal component (PC) regression and kernel machine (KM) regression further supported these findings and identified two additional ICRs (ICR_1177 and ICR_1180, mapping to DUX4L37 and *LINC01597,* respectively), which were also localized in the same region of chromosome 20 (Table 1). PC and KM regression -log10(p-value) show high correlation by Pearson’s correlation coefficient (Supplementary Figure S2). Furthermore, the methylation differences were largely directionally consistent across ICRs, with 76 out of 86 (88%) of CpG sites demonstrating higher methylation in the sustained obesity group compared to the normal weight. In the sensitivity analysis using the mean CpG methylation across each ICR, only ICR_1179 and ICR_1182 remained statistically significant (Supplementary Table S3).

### Imprinting control regions associated with intermittent childhood obesity

Comparison of normal weight and intermittent obesity (i.e., obese < 80% of the time, n = 108) using logistic regression yielded one significant CpG (q < 0.05) and significance at the ICR level (PC regression q < 0.05) for ICR_888 (Figure 1b, Supplementary Table S4). This ICR is within *NDN,* a known imprinted gene with paternal expression on 15q11.2 (78). *NDN* is associated with Prader-Willi syndrome, a disease that increases a child’s risk for obesity, largely due to hyperphagia (79). There was no overlap between the CpGs or ICRs found to be significant in the sustained obesity and intermittent obesity group analyses. In sensitivity analysis using the mean CpG methylation across each ICR, none of the ICRs remained statistically significant.

Association with putative ICR methylation are pronounced in offspring born to women with pre-pregnancy obesity

Because pre-pregnancy maternal obesity contributes substantially to childhood obesity and may represent a distinct etiology (80), we stratified by pre-pregnancy maternal obesity (BMI ≥ 30), and repeated the analysis for the normal weight versus sustained obesity groups. Despite a smaller size (n = 111 normal weight children and n = 33 sustained obesity children), these analyses yielded 54 significant ICRs with PC or KM regression q < 0.05 associated with sustained childhood obesity when the mothers were obese pre-pregnancy (Supplementary Table S5). Four of these ICRs overlap with the unstratified analysis (ICR_1182, ICR_1165, ICR_927, and ICR_1179). Among the ICRs that do not overlap with the unstratified analysis, four were within known imprinted domains: ICR_1205 in GNAS, ICR_475 in PEG10, ICR_721 in KCNQ1, and ICR_484 in SVOPL. No significant CpGs or ICRs with q < 0.05 were associated with sustained obesity in analysis stratified by child sex. Similarly, no CpGs or ICRs were significantly associated with intermittent obesity in analyses stratified by maternal obesity or child sex. Consistent with these findings, the sensitivity analysis using mean CpG methylation across each ICR also identified no significant associations with sustained or intermittent obesity in the stratified analyses.

### Evidence for novel putative ICR associated with childhood obesity on chromosome 20

Notably, a cluster of sustained obesity associated loci on chromosome 20q11.1–20q11.21—comprised of ICR_1165, ICR_1177, ICR_1179, ICR_1180, ICR_1181, and ICR_1182—suggests the presence of a previously unrecognized, contiguous regulatory domain rather than independent signals. Nineteen of the 20 CpG sites measured in ICR_1182 and neighboring ICR_1183 (mapping nearest pseudogene *CDC27P4*) showed increased methylation in the sustained obesity children compared to normal weight (Figure 2), suggesting that methylation at these CpG sites likely act together to modulate pathways contributing to childhood obesity risk.

**Figure 2.**
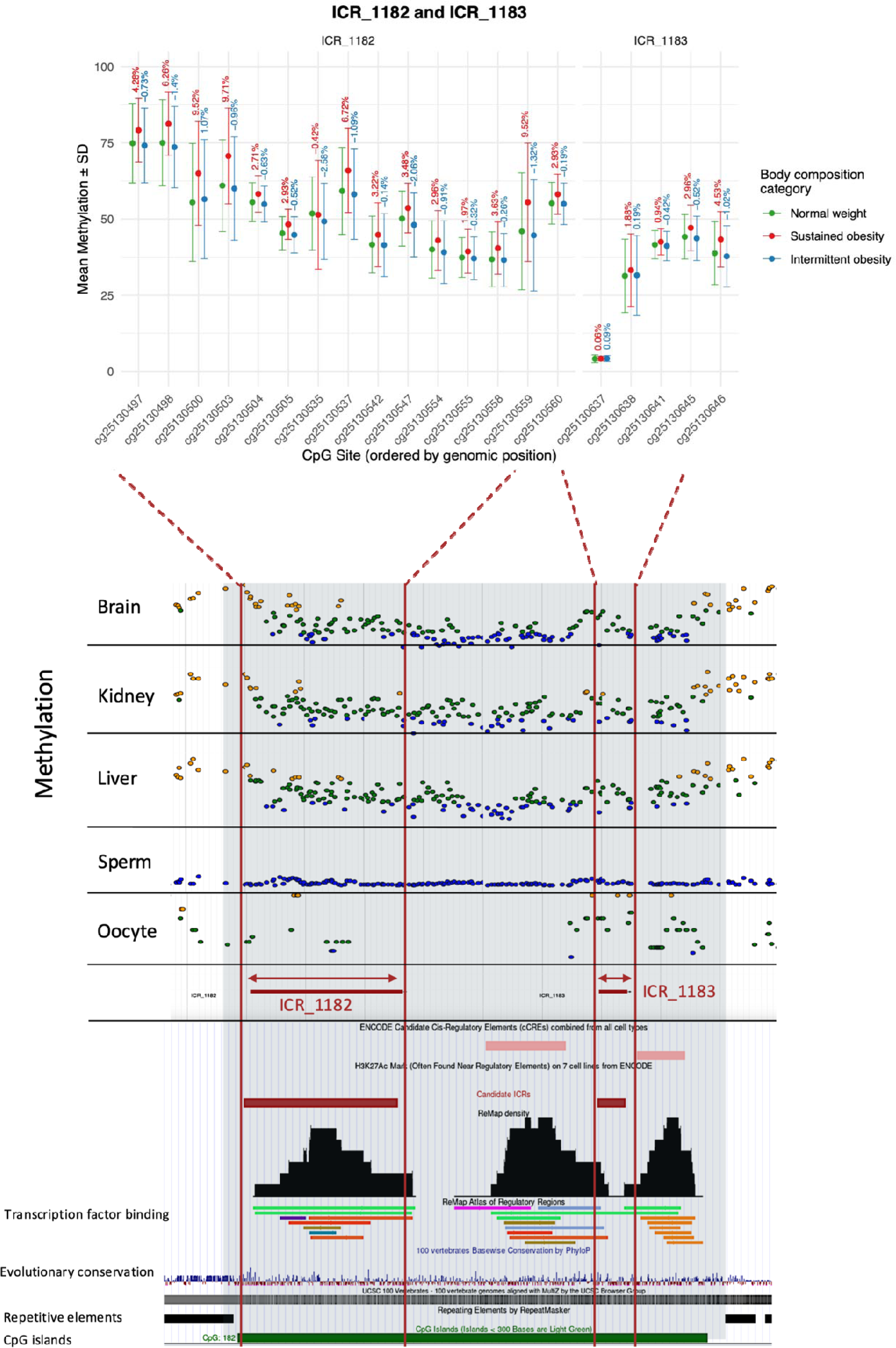
Methylation patterns and regulatory features of ICR_1182 and ICR_1183. (Top panel) Mean methylation levels at CpG sites in ICR_1182 and ICR_1183 across the three categories of body composition. Error bars correspond to the standard deviation of methylation percent within each body composition category. Labels indicate the percent-point difference in mean methylation (beta value) between normal weight and sustained obesity (red) or normal weight and intermittent obesity (blue) at that CpG site. Beta values were rescaled from 0-1 to 0-100 for interpretability. (Bottom panel) CpG site methylation in ICR_1182 and ICR_1183 from whole genome bisulfite sequencing in human fetal tissues. Methylation in germline (sperm and oocyte) and somatic (brain, kidney, and liver) tissues are colored orange for hypermethylation (> 65%), green for hemimethylation (35-65%), and blue for hypomethylation (< 35%) (https://humanicr.org/). Transcription factor binding, evolutionary conservation, repetitive elements, and CpG islands were identified by the UCSC Genome Browser. Transcription factor binding sites are color-coded as follows: RORC (green), NR3C1 (purple), ERG (bright red), SREBP2 (light red), BCOR (brown), MBD2 (teal), TRIM24 (pink), EZH2 (gray-blue), and CTCF (orange).

Multiple lines of evidence further support this locus as a candidate ICR, including germline methylation patterns consistent with allele-specific regulation and a stable ∼50% methylation profile across somatic tissues from all three germ layers (Figure 2, humanicr.org). In addition, the colocalization of these regions with transcription factor binding sites, including the important chromatin regulator CTCF, provides mechanistic support for their role in regulating parent-of-origin–specific epigenetic states. Similar associations were found with ICR_1179, ICR_1180, and ICR_1181, which are located within or proximal to the noncoding RNA gene *LINC01597*, supporting the possibility of coordinated regulation. Finally, we identified associations at ICR_1165 and ICR_1177, mapping to the pseudogenes *DUX4L34* and *DUX4L37*, respectively, within repetitive D4Z4 regions. While these "DUX4-like" sequences have primarily been studied in the context of muscular dystrophy and cancer (81), their identification here represents a novel link between these repetitive genomic elements and childhood obesity.

### Stability of ICRs from birth to late childhood/adolescence

A clinically important feature of ICR methylation signatures is their temporal stability across the life course, as methylation patterns that change substantially over time have limited utility for predicting later disease risk. Therefore, for ICRs significantly associated with sustained obesity in the unstratified analysis (by CpG logistic regression q < 0.05 or PC/KM regression q < 0.05), we evaluated the persistence of these methylation marks by calculating Pearson’s correlation coefficients and comparing group mean methylation levels in cord blood-derived DNA at birth and peripheral blood collected from the same children at ages 8-16 years (n = 86 normal weight and n = 14 sustained obesity children). We observed remarkably high correlations in ICR methylation between timepoints for the six ICRs located on chromosome 20. Pearson’s correlation coefficients ranged from 0.71–0.91 for the normal weight group, and 0.68–0.85 for the sustained obesity group (Table 2). ICR_927 did not show any correlation between ICR methylation measurements at birth and later childhood/adolescence. The ICR mean differences between the normal weight and sustained obesity children at birth, during later childhood/adolescence, and the changes between timepoints are summarized in Table 2. Most ICRs show group mean shifts of less than 1% between birth and later childhood/adolescence, and only ICR_1182 retained a PC regression-adjusted p-value below 0.05.

**Table 2.** Stability of methylation at ICRs between birth and later childhood/adolescence. Pearson’s correlation coefficient (r) between birth at age 0 and late childhood/adolescence at ages 8-16. Column A is the mean DNA methylation difference (sustained obesity – normal weight) from umbilical cord blood at birth, and Column B is the mean DNA methylation difference (sustained obesity – normal weight) from peripheral blood in late childhood/adolescence. Both timepoints used the same subset of children (n = 86 normal weight and n = 14 sustained obesity). All ICRs exhibited higher methylation in children with sustained obesity compared to normal weight at both timepoints. ICRs with less than a 1 percentage point change in methylation between the two timepoints are bolded in “Column B – Column A.” The final column reports PC regression adjusted p-values for the ICRs tested for an association with obesity later in life, with adjusted p < 0.05 bolded.

| ICR | Pearson's r –<br>normal weight | Pearson's r –<br>obese | **Column A**<br>Mean methylation<br>difference at birth<br>(obese – normal weight;<br>percentage points) | **Column B**<br>Mean methylation<br>difference in late<br>childhood/adolesce<br>nce (obese – normal<br>weight; percentage<br>points) | **Column B - Column A**<br>Change between late<br>childhood/adolescence and<br>birth (percentage points) | PC<br>Regression<br>FDR-adjusted p<br>for late<br>childhood/a<br>dolescence |
| --- | --- | --- | --- | --- | --- | --- |
| ICR_1165 | 0.82 | 0.73 | 2.03 | 1.73 | **-0.30** | 0.065 |
| ICR_1177 | 0.71 | 0.72 | 2.66 | 2.63 | **-0.03** | 0.135 |
| ICR_1179 | 0.85 | 0.70 | 3.75 | 4.54 | **0.79** | 0.255 |
| ICR_1180 | 0.91 | 0.85 | 4.59 | 4.05 | **-0.54** | 0.855 |
| ICR_1181 | 0.83 | 0.68 | 1.29 | 1.78 | **0.49** | 0.075 |
| ICR_1182 | 0.89 | 0.71 | 7.09 | 5.07 | -2.02 | **0.025** |
| ICR_927 | 0.17 | -0.02 | 1.52 | 1.01 | **-0.51** | 0.325 |

### Comparison with previous findings

Previous epigenetic studies have investigated the association of differential methylation with childhood obesity and identified plausible CpG sites using the Illumina Infinium HumanMethylation450 (450K) or EPIC BeadChip (EPIC) arrays, but replicating these findings has been challenging. Comparison of our findings with these prior studies (33,82–84) revealed limited overlap at the individual CpG-level, with concordance emerging primarily when considering genes mapping to associated CpGs or ICRs. Relaxing the statistical significance to include all CpGs and ICRs with q < 0.20 in the unstratified and stratified analyses to capture additional regions of potential relevance (Supplementary Tables S3-S6 and Supplementary Figures S3 and S4) revealed multiple genes annotated near significant or borderline significant ICRs overlapping with previous reports. Among these were 12 genes for which imprinting has been characterized (*BLCAP, NNAT, DLGAP2, GLI3, GNAS, KCNQ1, KCNQ1DN, MAGEL2, MEST, PEG3, PTPRN2*, and *SVOPL*; see geneimprint.com for a full list of imprinted genes for humans), and 5 putative novel ICRs located near or overlapping genes *ADARB2, CRYL1, OPCML, TBCD, and TRAPPC9* (Table 3). In total, 65 genes measured by the Human Imprintome array with PC or KM regression q < 0.20 in the unstratified or stratified analyses overlapped with genes reported as significant in previous studies (Supplementary Table S7). All previous BMI-based studies adjusted BMI for age and sex, consistent with our analytical approach.

**Table 3.**
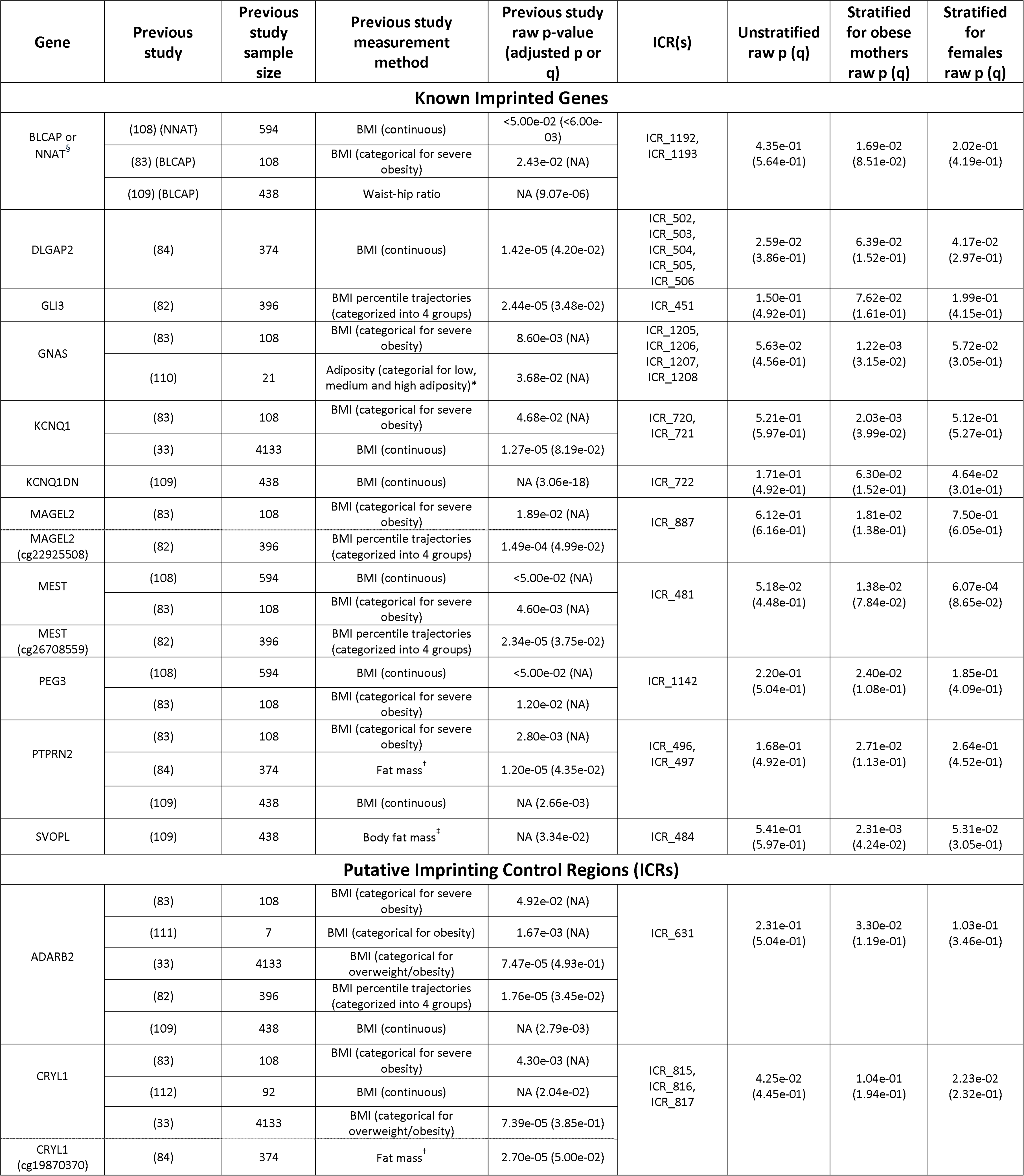
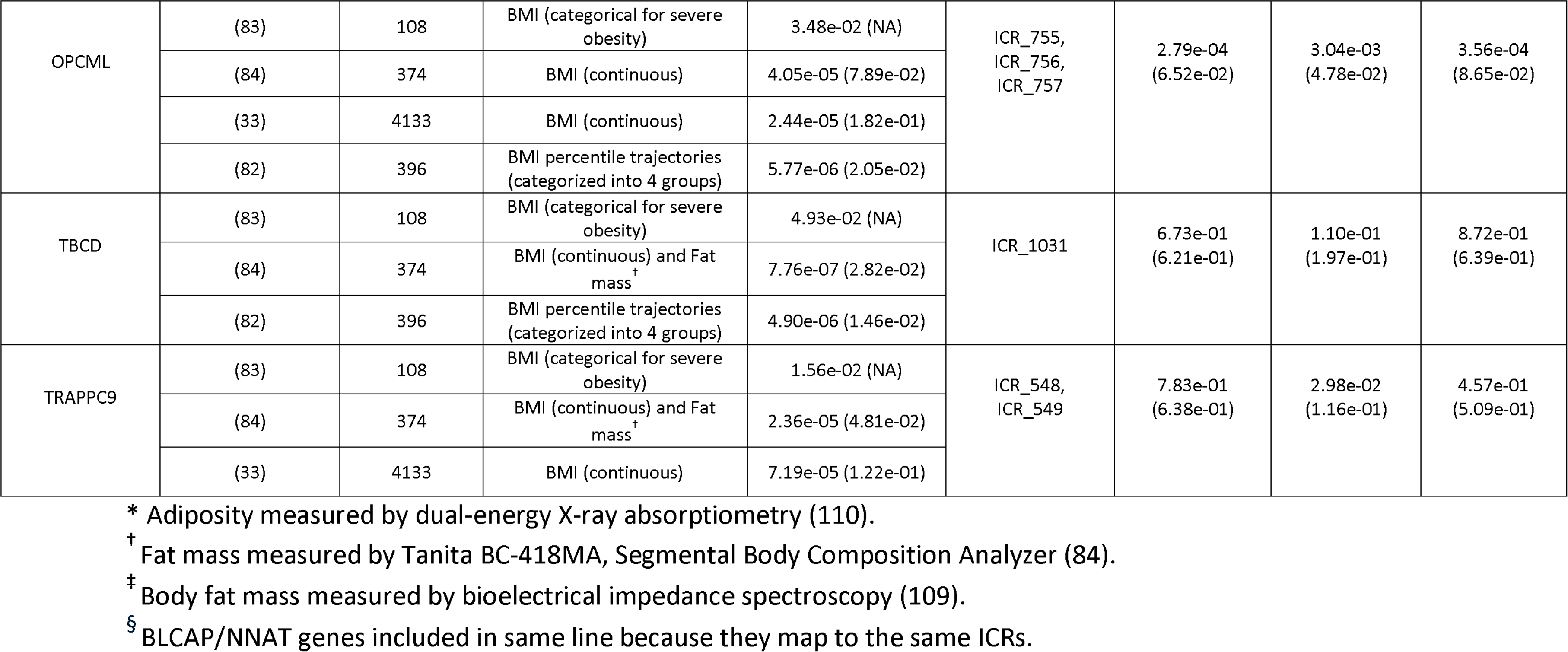
Comparison of ICR-associated genes identified in this study with genes previously associated with childhood obesity through differential methylation. All known imprinted genes identified as significant or borderline significant in our analysis and previous studies are shown in the table, along with the top five differentially methylated genes identified in at least three previous studies. The results from previous studies are reported with their p-value and multiple-testing adjusted p-value, or q-value, where applicable. The p- and q-values refer to PC or KM regression values for the entire ICR (if both are significant the lower value is shown; similarly, if multiple ICRs map to the same gene, then the lowest value is shown). If the same CpG appeared in our analysis and previous studies, the CpG ID is noted in parentheses next to the gene name.

### Functional pathway analysis

The Ingenuity Pathway Analysis (IPA) was performed on the 58 genes mapping to a CpG or ICR with q < 0.05, out of a background of 899 total genes located near CpGs or ICRs measured on the array. The top 20 most significant pathways identified by IPA are presented in Table 4. Overall pathway analysis of ICR-associated genes provides strong biological support that both putative and established imprinting control regions identified in this study are relevant to obesity and metabolic dysfunction detectable at birth. Several significant ICRs (e.g., ICR_1359, ICR_1205, and ICR_446) map to genes such as *MAPK1*, *GNAS*, and *CREB5*, which recur across multiple enriched pathways involved in energy homeostasis, adipose tissue biology, and metabolic signaling. Notably, pathways such as white adipose tissue browning and PFKFB4 signaling are directly linked to metabolic regulation and energy expenditure, while inflammatory and immune-related pathways (e.g., IL-4, IL-10, IL-13, and macrophage alternative activation signaling) highlight early-life immune-metabolic crosstalk known to influence obesity risk (85). In addition, enrichment of neuroendocrine and neuronal signaling pathways—including corticotropin-releasing hormone signaling, glutamatergic receptor signaling, and sleep-related pathways—suggests involvement of central regulatory mechanisms governing appetite, stress response, and circadian metabolism (86). The repeated involvement of key regulatory genes across these diverse pathways suggests that methylation differences at these ICRs represent processes underlying metabolic programming. These genes and pathways implicate obesity development through diverse metabolic, neural, hormonal, and inflammatory mechanisms.

**Table 4.** Functional analysis of significant ICR-associated genes using Ingenuity Pathway Analysis. The top 20 most significant pathways by IPA p-value are displayed, alongside the input genes contributing to each pathway and the ICRs nearest those genes.

| Pathway | Genes associated with pathway | Significant ICR(s) mapping to genes |
| --- | --- | --- |
| Transcriptional Regulation by NPAS4 | *MAPK1, RBFOX3, RET* | ICR_1359, ICR_1027, ICR_654 |
| PFKFB4 Signaling Pathway | *CREB5, GNAS, MAPK1* | ICR_446, ICR_1205, ICR_1359 |
| White Adipose Tissue Browning Pathway | *CACNG2, CREB5, GNAS, MAPK1, NDN* | ICR_1369, ICR_446, ICR_1205, ICR_1359, ICR_888 |
| IL-4 Signaling | *COL5A1, CREB5, IL4R, MAPK1* | ICR_621, ICR_446, ICR_933, ICR_1359 |
| Glutaminergic Receptor Signaling Pathway (Enhanced) | *CACNG2, CREB5, DGKI, GNAS, MAPK1* | ICR_1369, ICR_446, ICR_483, ICR_1205, ICR_1359 |
| Ephrin Receptor Signaling | *CREB5, GNAS, MAPK1, RAPGEF1* | ICR_446, ICR_1205, ICR_1359, ICR_618 |
| IL-10 Signaling | *CREB5, IL4R, MAPK1* | ICR_446, ICR_933, ICR_1359 |
| Melanocyte Development and Pigmentation Signaling | *CREB5, GNAS, MAPK1* | ICR_446, ICR_1205, ICR_1359 |
| Macrophage Alternative Activation Signaling Pathway | *CREB5, IL4R, MAPK1* | ICR_446, ICR_933, ICR_1359 |
| Sleep NREM Signaling Pathway | *CREB5, GNAS, MAPK1* | ICR_446, ICR_1205, ICR_1359 |
| CGRP in Inflammation and Nociception Signaling Pathway | *CREB5, GNAS, MAPK1* | ICR_446, ICR_1205, ICR_1359 |
| Docosahexaenoic Acid (DHA) Signaling | *CREB5, GNAS, MAPK1* | ICR_446, ICR_1205, ICR_1359 |
| Corticotropin Releasing Hormone Signaling | *CACNG2, CREB5, GNAS, MAPK1* | ICR_1369, ICR_446, ICR_1205, ICR_1359 |
| IL-13 Signaling Pathway | *IL4R, MAPK1* | ICR_933, ICR_1359 |
| Signaling by PDGF | *COL5A1, RAPGEF1* | ICR_621, ICR_618 |
| Interleukin-4 and Interleukin-13 signaling | *IL4R, NDN* | ICR_933, ICR_888 |
| Sleep REM Signaling Pathway | *CREB5, MAPK1* | ICR_446, ICR_1359 |
| Role of IL-17F in Allergic Inflammatory Airway Diseases | *CREB5, MAPK1* | ICR_446, ICR_1359 |
| GDNF Family Ligand-Receptor Interactions | *MAPK1, RET* | ICR_1359, ICR_654 |
| FGF Signaling | *CREB5, MAPK1* | ICR_446, ICR_1359 |

## Discussion

Using the novel Human Imprintome Array, we tested whether differential methylation at imprinting control regions (ICRs) at birth is associated with longitudinal childhood obesity and if these methylation differences persist into later childhood. Eight ICRs, most located in the long arm of chromosome 20, were significant (q < 0.05) in the unstratified analyses. These include ICR_1182, ICR_1179, ICR_1181, ICR1177, ICR_1180, ICR_1165, and ICR_927 (mapping to *CDC27P4, LINC01597, DUX4L37, DUX4L34*, and *ZNF597/NAA60*) which were associated with sustained obesity, and ICR_888 (mapping to NDN) which was associated with intermittent obesity. While two (ICR_927 and ICR_888) are within previously described imprinted domains related to metabolic and imprinting disorders, the majority (six) represent putative novel regions. Remarkably, in analysis stratified for children born to mothers with pre-pregnancy obesity, a total of 54 ICRs were associated with sustained obesity. Four of these overlapped with the unstratified sustained obesity analysis: ICR_1182 (*CDC27P4*), ICR_1179 (*LINC01597*), ICR_1165 (*DUX4L34*), and ICR_927 (*ZNF597/NAA60*). Although not discernable in sensitivity analyses that used the simple arithmetic mean of CpG methylation to define the ICR, highlighting the benefit of these ICR-level analytical approaches, these findings warrant replication in larger studies, and experimental validation. Nonetheless, this predominance of previously unreported ICRs suggests that the findings may meaningfully expand the current catalog of imprinted loci, particularly in the context of metabolic regulation. Effect direction and magnitude for sustained obesity remained consistent over time, supporting stability of ICR methylation from birth through at least adolescence. Furthermore, regardless of whether the children were normal weight or sustained obesity, all ICRs on chromosome 20 displayed stability between birth and late childhood/adolescence.

Of the seven sustained obesity-associated ICRs, three (ICR_1179, ICR_1180, and ICR_1181) map to the *LINC01597* RNA gene. With boundaries of ICRs not yet experimentally defined, it is possible that these three separate ICRs constitute a single region that regulates the imprinting of *LINC01597*. Six significant ICRs are located on chromosome 20q11.1-20q11.21, implicating this region of the genome in epigenetic regulation of obesity. ICR_1182 has compelling evidence to be considered a bona fide ICR and even retained adjusted p < 0.05 in the stability analysis. ICR_1182 is likely part of a larger single regulatory region of ∼2,100 bp, in combination with ICR_1183 (with both ICRs mapping closest to the *CDC27P4* pseudogene). As seen in Figure 2, across the ∼800 bp between the two ICRs, methylation in the somatic brain, liver, and kidney tissues (representing the three germ layers) falls just outside the strict limits required by the original candidate ICR definition (54). Also notable is the CpG island spanning both ICRs and three clusters of transcription factor binding sites, one neatly aligning with ICR_1182 and two flanking ICR_1183, strongly suggesting a regulatory role in modulating transcription factor activity and nearby gene expression that may contribute to sustained obesity. Conversely, the large regions in *LINC01597* or *CDC27P4* may represent correlated regions of systemic interindividual variation (CoRSIVs) rather than ICRs. Indeed, the region encompassing ICR_1159–ICR_1191 has been found to be densely populated with highly correlated CoRSIVs, yet sparsely populated with genes (87). While both ICRs and CoRSIVs show high methylation agreement across tissues and are affected by periconceptional environmental factors, CoRSIVs are a broader class of epigenetic regulation that display greater methylation variation between individuals, and are not defined by parent-of-origin-specific methylation (87,88). Given these features, any mechanistic and exposure studies based on the identification of ICRs on chromosome 20q11.1-20q11.21 in connection to childhood obesity should consider this entire region.

Because the chromosome 20q11.1-20q11.21 region has not been previously implicated in childhood obesity, replication in independent cohorts will be an important next step to establish the robustness and reproducibility of these findings. In addition, long read sequencing platforms such as Oxford Nanopore Technologies, could be used to evaluate allele-specific methylation across this region. Unlike short-read sequencing, long-read sequencing captures multiple heterozygous variants within individual DNA molecules, enabling haplotype phasing without requiring parental genotypes. These analyses could confirm whether this region exhibits the allele-specific methylation expected of a bona fide ICR, refine the boundaries of the novel putative ICRs, and determine whether adjacent candidate ICRs represent distinct regulatory elements or components of a larger imprinting domain.

Our findings align with imprinting biology and metabolic diseases. The established imprinted gene *ZNF597* has been linked to a clinical case phenotypically similar to Silver-Russell Syndrome (SRS) (89), a congenital imprinting disorder which regulate genes critical for growth and metabolism (90). Aberrant methylation in *ZNF597* for patients with obesity and type 2 diabetes further provides a link between *ZNF597* and metabolic dysfunction (91). The ICRs identified in children born to mothers with pre-pregnancy obesity included regions in several established imprinted genes including ICR_1205 (*GNAS*), ICR_484 (*SVOPL*), ICR_475 (*PEG10*), and ICR_721 (*KCNQ1*). *GNAS* is a well-characterized imprinted gene with maternal or paternal expression, depending on the transcript isoform (92,93). *GNAS* is known to be involved in obesity through genetic and epigenetic influences. Maternal *GNAS* mutations can cause pseudohypoparathyroidism type 1A (PHP1A), a disease that often coincides with early-onset obesity (94). Additionally, differential methylation in CpG sites of SVOPL, a maternally expressed gene (95), has been found in conjunction with GNAS methylation abnormalities in PHP patients (96). *PEG10* is a paternally expressed gene involved in early adipocyte differentiation in mice and has previously been associated with increased methylation in offspring of either obese mothers or obese fathers (31,97). The methylation levels of *KCNQ1*, a maternally expressed gene, have been previously shown to differ in obese individuals (98). These observations support the biological relevance of the identified ICRs and further implicate imprinted gene regulation as a potentially important contributor to early-life metabolic programming associated with obesity.

Interestingly, the ICRs found to be significant for unstratified intermittent obesity did not overlap with those in the unstratified sustained obesity analysis, possibly indicating different etiology of these phenotypes. While intermittent obesity is not a standard clinical term, it describes children whose BMI fluctuates over their childhood such that they cannot be characterized as sustained obesity or normal weight. Five CpGs map to ICR_888 within the NDN gene and all show hypomethylation in the intermittent obesity group. The NDN gene is part of the melanoma antigen (MAGE) family which includes *MAGEL2* (99), an imprinted gene with paternal expression in the Prader-Willi Syndrome (PWS) region of the genome. *MAGEL2* has been associated with obesity in previous studies (Table 3) (82,83), providing a potential connection between *NDN* and childhood obesity. Furthermore, *MAGEL2* and *NDN* were found to regulate leptin receptor activity in obese mice, implicating these genes in the development of obesity in individuals with PWS (100).

The IPA functional analysis was primarily driven by the genes identified in the maternal obesity-stratified analysis, because this analysis yielded the largest number of significant ICRs. The top 20 pathways returned are involved in a range of biological processes, including metabolic pathways (e.g. white adipose tissue browning and PFKFB4 signaling), neurocognitive pathways (e.g. transcriptional regulation by NPAS4 and glutaminergic receptor signaling), and immune pathways (e.g. IL-10 and macrophage signaling). These top 20 pathways are driven by only 11 of the significant genes (Table 4), suggesting that abnormal methylation in relatively few genes can influence multiple interconnected downstream pathways. Browning of white adipose tissue is protective against obesity (101), and thus disruption in this pathway may promote obesity development. IPA yielded several neural-related pathways, consistent with previously reported connection between neural function and obesity (102). For example, NPAS4 was recently implicated in feeding behaviors, with *Npas4* knockout mice exhibiting decreased nutrient intake and lower bodyweight (103). The FGF (fibroblast growth factor) signaling pathway plays a crucial role in growth and development. FGF signaling is active during early embryonic development and involved in adult metabolism (104), suggesting that changes in this pathway may affect obesity prognosis throughout an individual’s life. Altogether, these pathways show that even at birth, the methylation profiles of infants born to mothers with pre-pregnancy obesity are already primed for obesogenic risk through diverse mechanisms and interrelated pathways.

Studies relying largely on 450K/850K methylation arrays, which interrogate <5% of genomic CpGs, generally show greater reproducibility at the regional or gene level than at individual CpG sites. Illustrating this limitation, despite overlapping cohorts—including our cohort (NEST) and the European Childhood Obesity Project (CHOP)—we identified no CpGs in common with the 23-cohort meta-analysis by *Vehmeijer et al.* (33). Similarly, *Rzehak et al.* (84), analyzing CHOP, identified only two CpG sites that overlapped with *Vehmeijer et al.*, emphasizing the limited reproducibility of individual CpG site associations across studies. In contrast, Table 3 demonstrates considerable overlap at the gene-level between our study and prior studies, highlighting the value of examining regions of differential methylation. By focusing on ICRs rather than individual CpG sites, the imprintome-wide framework complements traditional epigenome-wide association studies (EWAS) by evaluating biologically defined regulatory domains that coordinate the expression of multiple genes. This region-based approach may improve biological interpretation, facilitate comparisons across studies, and enhance reproducibility by reducing reliance on isolated CpG site associations that can vary across cohorts. Future obesity epigenetic studies should evaluate whether EWAS-associated CpGs localize within known or putative ICRs or occur near genes regulated by these domains, as this regulatory context may provide additional mechanistic insight. Conversely, conventional EWAS can identify obesity-associated loci outside the imprintome. Together, these complementary approaches provide a more comprehensive understanding of the epigenetic mechanisms contributing to childhood obesity.

Many identified associations that align with previous findings are only borderline significant in our study with the less-stringent q-value threshold of < 0.20. Notably, *OPCML* showed borderline significance, aligning with four prior studies linking *OPCML* to childhood obesity. Independent evidence further implicates *OPCML* in metabolic phenotypes, including visceral fat distribution in adult women and glycemic control in children with nonalcoholic fatty liver disease (105,106). Similarly, we found borderline significance in girls at the paternally expressed gene MEST (ICR_481; Supplementary Table S6), consistent with female-specific associations reported previously across multiple developmental stages (82). Some previously reported genes (e.g. IGF2R) were not identified in our analysis, likely due to differences in array coverage. The nearest CpG site to IGF2R tested on the Human Imprintome Array is over 23,000 base pairs from previously reported sites, indicating that CpGs on 450K/EPIC arrays do not always capture ICR-related regions.

There are several limitations to this study that should be considered and provide opportunities for future research. While the Human Imprintome Array is able to capture many more CpGs within ICRs than previously developed arrays, only 1,088 of the 1,488 putative ICRs (73.1%) are represented due to exclusion of low-quality probes during development of the array (55). After the probe filtering step, 1,022 of the 1,088 ICRs on the array (94%) remained in this analysis, with some ICRs only represented by a single CpG site, primarily due to limitations in array design. Future investigation will require complementary deeper sequencing using technologies that capture longer reads to improve ICR boundaries and confirm their role in the regulation of imprinted genes. In this study, genetic sequencing data were not available and, therefore, we could not evaluate the contribution of underlying genetic variation to DNA methylation, which has been shown to influence methylation at ICRs (107). Future studies integrating DNA methylation with whole-genome sequencing data will be important for disentangling genetic and epigenetic effects at these putative ICRs and clarifying their role in childhood obesity.

Whole-genome sequencing is particularly advantageous because many ICRs reside within intronic or other regulatory regions outside protein-coding exons. Such studies could identify methylation quantitative trait loci (meQTLs), CpG-SNPs, or genetic variants that alter regulatory protein binding sites, thereby helping to determine whether the observed associations between ICR methylation and childhood obesity are independent of underlying genetic variation.

BMI is the most utilized measurement of obesity, but it approximates adiposity due to inability to separate muscle mass and fat mass. Other measurements, such as waist circumference, skinfold thickness, dual-energy X-ray absorptiometry, and bioelectrical impedance, may predict adiposity with greater accuracy or provide additional adiposity features such as abdominal fat. Nonetheless, the use of BMI in this analysis remains advantageous due to its translatability to other studies that have examined DNA methylation in childhood obesity. Anthropometric measures are easy to obtain during a routine doctor’s visit, and with increased data, investigating differential methylation in ICRs and its association with these measures of adiposity should be included in next steps. Repeated measures of metabolic markers (such as cholesterol, triglycerides, and glucose) are likely to lead to a better understanding of the role of differential methylation in ICRs in early detection of obesity and metabolic syndrome in children.

## Conclusion

In summary, we provide the first evidence that methylation changes on chromosome 20 at novel putative ICRs–which are established in the gametes–are associated with childhood obesity. These patterns persist into late childhood and early adolescence. Larger studies followed by experimental evidence for promising targets are required to determine the exact boundaries of these regions and confirm their dysregulation in obese children. Future work should also assess these findings in independent populations and across additional obesity-related endpoints, including obesity-related metabolic markers, to determine whether these candidate biomarkers can predict epigenetic predisposition to obesity at birth.

## Acknowledgements

The authors acknowledge the contributions of all Newborn Epigenetics STudy (NEST) participants and their families. The authors acknowledge the use of AI in this manuscript.

Microsoft Copilot (accessed 2025-2026) and ChatGPT by OpenAI (GPT-4 and later models; accessed 2025-2026) were used in refinement of scientific writing and coding assistance. Consensus AI (accessed 2025-2026) was used to search for published scientific literature. The authors are responsible for all analyses, interpretations, and conclusions.

## Data Availability

Data supporting the findings of this study have been deposited in the NCBI Gene Expression Omnibus (GEO) repository under accession number GSE334366. Upon publication, the data may be found at https://www.ncbi.nlm.nih.gov/geo/query/acc.cgi?acc=GSE334366.

## Code Availability

The analyses supporting these findings were implemented in R using the TruDiagnostic Human Imprintome Array (tdhia) package (71). To facilitate reproducibility, we have provided the code for downloading the data from GEO, processing the data, and conducting the analytical workflow in the publicly accessible hoyo_published_analyses GitHub repository.

## Ethics approval and consent to participate

This study was conducted in accordance with the principles stated in the Declaration of Helsinki and was approved by the Institutional Review Board of Duke University (Pro00014548). Written informed consent was obtained from mothers for their participation and their child’s participation.

